# Exploring the role of Large Language Models (LLMs) in hematology: a systematic review of applications, benefits, and limitations

**DOI:** 10.1101/2024.04.26.24306358

**Authors:** Aya Mudrik, Girish N Nadkarni, Orly Efros, Benjamin S Glicksberg, Eyal Klang, Shelly Soffer

## Abstract

**Rationale and Objectives:** Large Language Models (LLMs) have the potential to enhance medical training, education, and diagnosis. However, since these models were not originally designed for medical purposes, there are concerns regarding their reliability and safety in clinical settings. This review systematically assesses the utility, advantages, and potential risks of employing LLMs in the field of hematology.

**Materials and Methods:** We searched PubMed, Web of Science, and Scopus databases for original publications on LLMs application in hematology. We limited the search to articles published in English from December 01 2022 to March 25, 2024, coinciding with the introduction of ChatGPT. To evaluate the risk of bias, we used the adapted version of the Quality Assessment of Diagnostic Accuracy Studies criteria (QUADAS-2).

**Results:** Eleven studies fulfilled the eligibility criteria. The studies varied in their goals and methods, covering medical education, diagnosis, and clinical practice. GPT-3.5 and GPT-4’s demonstrated superior performance in diagnostic tasks and medical information propagation compared to other models like Google’s Bard (currently called Gemini). GPT-4 demonstrated particularly high accuracy in tasks such as interpreting hematology cases and diagnosing hemoglobinopathy, with performance metrics of 76% diagnostic accuracy and 88% accuracy in identifying normal blood cells. However, the study also revealed discrepancies in model consistency and the accuracy of provided references, indicating variability in their reliability.

**Conclusion:** While LLMs present significant opportunities for advancing clinical hematology, their incorporation into medical practice requires careful evaluation of their benefits and limitations.

## INTRODUCTION

LLMs such as OpenAI’s ChatGPT, Google’s Gemini, and Anthropic’s Claude, are reshaping the field of text generation. Notably, ChatGPT-3.5 alone boasts more than 100 million monthly active users.^1^

LLMs are increasingly used in clinical research and practice. They serve various purposes such as simplifying complex medical terminology for patient education.^2^ They also support physician training and academic learning through interactive tools^3^ and enhance diagnostic accuracy by creating predictive models from Electronic Health Records (EHR) data for patient outcomes.^4^ In hematology, a field with strict protocols and standards, the potential of LLMs goes beyond simple text interactions. They could significantly reshape the field.^5^

However, adopting LLMs, which were not initially designed for medical purposes, raises concerns regarding their reliability, including challenges in contextual understanding and interpretability, biases in the training data, and ethical and regulatory issues. Doubts also persist about the dependability of their outputs for making clinical decisions.^6^ ^7^ As LLMs become more common in healthcare, the necessity to test their applications increases. This review evaluates the application of LLMs in the field of hematology, systematically assessing their benefits, limitations, and potential risks in medical training, education, and diagnosis. It includes a thorough examination of studies from major databases, focusing on their use since the introduction of ChatGPT. The review guides us toward cautious yet optimistic integration of LLMs into clinical hematology practice.

### Tokens

Tokens are the basic units of data processed by LLMs. In the context of text, a token can be a word, part of a word or a character.^29^

### Prompt

In the context of AI, a "prompt" refers to the input given to a language model to initiate and guide its output generation.^29^

### Autoregression

Autoregression in AI refers to the process where a model predicts the next word or sequence based on the previous inputs. It operates by evaluating the probabilities of various possible continuations and selecting the most likely next element in the sequence.^29^

## METHODS

### Search Strategy

A systematic review was conducted according to the Preferred Reporting Items for Systematic Reviews and Meta-Analyses statement (PRISMA),^8^ relevant guidelines from the diagnostic test accuracy extension,^9^ and the recommendations for systematic reviews of prediction models (CHARMS checklist).^10^

A systematic search of the published literature was conducted on March 25, 2024. PubMed, Web of Science, and Scopus were used as databases.

Our search strategy targeted original studies at the intersection of LLMs and hematology, employing a set of search terms relevant to both fields. The detailed search strategy is outlined in the **Supplementary Materials** ("Detailed Search Terms").

We limited the search to articles published in English after December 1, 2022, to align with the introduction of ChatGPT, marking the first widespread release of LLMs.

Peer-reviewed original publications on the subject of LLMs applications in Hematology were included. We excluded articles that were not related to applications of LLMs in hematology, articles that were not original, and conference abstracts. To ensure that we did not inadvertently exclude relevant articles, we searched the bibliographies of the articles included in our study.

The study is registered with PROSPERO (CRD42024525241).

### Study Selection

Two reviewers (AM and SS) independently screened the titles and abstracts to determine whether the studies met the inclusion criteria. In unclear cases, the full-text article was reviewed. Disagreements were adjudicated by a third reviewer (EK). The two authors (AM and SS) independently assessed the full texts of the included articles.

### Data Extraction

Data from all included studies was collected into a standardized data extraction sheet. Data included publication year, LLM model types, objective, sample size, main findings, and limitations.

### Quality Assessment and Risk of Bias

To evaluate the risk of bias, we used the adapted version of the Quality Assessment of Diagnostic Accuracy Studies criteria (QUADAS-2).^11^

### Data Synthesis

We conducted a narrative synthesis of the findings from the included studies. Due to the heterogeneity in the study designs and outcomes, a meta-analysis was not implemented. Instead, we focused on summarizing the applications, benefits, and limitations of LLMs in Hematology as reported in the studies.

## RESULTS

A total of 325 articles were retrieved in the initial search. After exclusion **(Supplementary** Figure 1), 11 studies evaluating the application of LLMs in hematology were included.

The included studies were diverse in their objectives and methodologies and covered various aspects of hematology including lymphoma, transfusion medicine, hemophilia, hemoglobinopathies, and stem cell transplant. Categories included medical education, diagnosis, and clinical practice **(Figure3)**.

All studies were evaluated for risk of bias and applicability using the QUADAS-2 tool **(Supplementary Table 1)**. Generally, the studies demonstrated a low risk of bias across the criteria of index test, reference standard, and flow and timing. In some studies, the composition of the sample (patient selection criteria) presented an intermediate risk of bias. For example, in their article, Kumari A. et al.^12^ wrote 50 hematologic questions that were posed to the LLMs. The authors chose which questions to ask, which might have influenced the outcomes of the study.

The characteristics of the studies are presented in **Table 1**. Objectives, reference standards, sample sizes, and main findings are presented in **Table 2**. The strengths and limitations of LLMs as presented in the included studies are summarized in **Table 3**. **In the following section, we detail the included studies separated by theme.**

**Table 1.** Details about the reviewed articles.

| Group | Title | First Author | Journal/Book | Year |
| --- | --- | --- | --- | --- |
| Medical Education | Using A Google Web Search Analysis to Assess the Utility of ChatGPT in Stem Cell Therapy | Chen L<sup>13</sup> | Stem Cells Translational Medicine | 2024 |
|  | Evaluation of OpenAI's large language model as a new tool for writing papers in the field of thrombosis and hemostasis | Klang E<sup>17</sup> | Journal of Thrombosis and Haemostasis | 2023 |
|  | Consulting the Digital Doctor: Google Versus ChatGPT as Sources of Information on Breast Implant-Associated Anaplastic Large Cell Lymphoma and Breast Implant Illness | Liu HY<sup>14</sup> | Aesthetic Plastic Surgery | 2023 |
|  | How good does ChatGPT answer frequently asked questions about haemophilia? | Van de Wyngaert C<sup>15</sup> | Haemophilia | 2023 |
| Medical Diagnosis | Large Language Models in Hematology Case Solving: A Comparative Study of ChatGPT-3.5, Google Bard, and Microsoft Bing | Kumari A<sup>12</sup> | The Cureus Journal of Medical Science | 2023 |
|  | Predicting hemoglobinopathies using ChatGPT | Kurstjens S<sup>18</sup> | Clinical Chemistry and Laboratory Medicine | 2023 |
|  | ChatGPT's innovative application in blood morphology recognition | Yang WH<sup>19</sup> | Journal of the Chinese Medical Association | 2024 |
| Clinical Practice | Evaluating the performance of large language models in haematopoietic stem cell transplantation decision-making | Civettini I<sup>22</sup> | British Journal of Haematology | 2023 |
|  | Thromboembolic prophylaxis in spine surgery: an analysis of ChatGPT recommendations | Duey AH<sup>20</sup> | The Spine Journal | 2023 |
|  | Would doctors dream of electric blood bankers? Large language model-based artificial intelligence performs well in many aspects of transfusion medicine | Hurley NC<sup>16</sup> | Transfusion | 2023 |
|  | Battle of the (Chat)Bots: Comparing Large Language Models to Practice Guidelines for Transfusion-Associated Graft-Versus-Host Disease Prevention | Stephens LD<sup>21</sup> | Transfusion Medicine Reviews | 2023 |

**Table 2.** A summary of the reviewed articles.

| Group | First Author | Model Used | Objective | Reference Standard | Sample Size | Main Findings |
| --- | --- | --- | --- | --- | --- | --- |
| Medical Education | Chen L<sup>13</sup> | ChatGPT3.5 | Compared the effectiveness of ChatGPT to Google in answering medical questions related to stem cell therapy | Google search | 20 questions | Compared to Google, ChatGPT exhibits stronger capabilities in promoting awareness of stem cell therapy |
|  | Hurley NC<sup>16</sup> | ChatGPT3.5<br>ChatGPT4<br>Google Bard | Testing the explicit transfusion medicine knowledge of different LLMs | The official exam's results | 20 questions | ChatGPT3.5- 44%,<br>ChatGPT4- 87%<br>Bard 54% correct answers |
|  | Klang E<sup>17</sup> | ChatGPT3.5 | Explore the potential benefits and restrictions of using chatGPT for academic writing | A hematology expert specialist in thrombosis and hemostasis | Three commentaries | There are many limitations in the commentaries generated by ChatGPT which highlighted the importance of human judgment and medical expertise in the writing of academic papers |
|  | Liu HY<sup>14</sup> | ChatGPT3.5 | Assess the quality of ChatGPT as a potential source of patient education | Five breast plastic surgeons | 10 questions | (from scale of 1-5)<br>ChatGPT: 4.18±1.04<br>Google: 2.72±1.44 |
|  | Van de Wyngaert C<sup>15</sup> | ChatGPT | Assess the quality and validity of ChatGPT's answers to frequently asked questions about haemophilia | Twenty hemophilia experts | 15 questions | (from scale of 1-5)<br>ChatGPT:<br>Scientific validity- 4.04<br>Exhaustivity- 3.94<br>Relevance- 4.01<br>Understandability- 4.08<br>Websites:<br>Scientific validity- 3.55<br>Exhaustivity- 2.90<br>Relevance- 3.30<br>Understandability- 3.62 |
| Medical Diagnosis | Kumari A<sup>12</sup> | ChatGPT3.5<br>Google Bard<br>Microsoft Bing | Exploring the capability of LLMs in solving hematology cases and conducting a comparative analysis of three LLMs | Three raters with expertise in hematology and medical education | 50 questions | (from scale of 1-5)<br>ChatGPT (3.15±1.19)<br>Bard (2.23±1.17),<br>Bing (1.98±1.01). |
|  | Kurstjens S<sup>18</sup> | ChatGPT3.5<br>ChatGPT4 | Investigate the capability of ChatGPT to diagnose hemoglobinopathy by interpreting the patient's laboratory results of CBC and ferritin values | Medical diagnosis | 59 patients | ChatGPT-3.5:<br>Accuracy-53%,<br>True Positive-75%,<br>True Negative-31%,<br>ChatGPT-4.0:<br>Accuracy-76%,<br>True Positive-97%,<br>True Negative-56%, |
|  | Yang WH<sup>19</sup> | ChatGPT4 | Investigate the potential of ChatGPT-4 to assist with blood morphology identification | Public image database of American Society of Hematology | 38 JPEG images depicting 44 features | ChatGPT4: 69%<br>Manual Identification: 71% |
| Clinical Practice | Civettini I<sup>22</sup> | ChatGPT4<br>PaLm2<br>Llama2 13b<br>Llama2 70b | Assess the capabilities of LLMs in making complex decisions in haematopoietic stem cell transplantation | Six bone marrow transplant specialists | 150 questions | LLMs- 58.8%<br>Residents- 76.5% |
|  | Duey AH<sup>20</sup> | ChatGPT3.5<br>ChatGPT4 | Evaluate the recommendations given by ChatGPT for thromboembolic prophylaxis in spine surgery | North American Spine Society (NASS) clinical guidelines | 12 questions | ChatGPT-3.5:<br>Accuracy-33%<br>Over-Conclusive-50%,<br>Supplemental-67%<br>Incomplete-33%.<br>ChatGPT-4.0:<br>Accuracy-92%,<br>Over-Conclusive-8%<br>Supplemental-92%<br>Incomplete-33%. |
|  | Hurley NC<sup>16</sup> | ChatGPT3.5<br>ChatGPT4<br>Google Bard | Evaluate the capabilities of different LLMs in answering common questions in clinical transfusion practice | Grading rubric developed by the authors | 12 questions | ChatGPT3.5:<br>Implicit questions- 17%<br>Explicit questions- 83%<br>ChatGPT4:<br>Implicit questions- 83%<br>Explicit questions- 100%<br>Bard:<br>Implicit questions- 33%<br>Explicit questions- 33% |
|  | Hurley NC<sup>16</sup> | ChatGPT3.5<br>ChatGPT4<br>Google Bard | Evaluate the recommendations of different LLMs regarding the necessity for red blood cell transfusion, based on short case presentations | AABB recommendations and a restrictive transfusion strategy | 44 questions | ChatGPT3.5- 63.6%,<br>ChatGPT4- 91%<br>Bard 45.5% |
|  | Stephens LD<sup>21</sup> | ChatGPT3.5<br>ChatGPT4<br>Google Bard<br>Microsoft Bing | Evaluate the quality of the indications for irradiation of blood component for TA-GVHD prevention, provided by LLMs and conducting a comparative analysis of four LLMs | 4 transfusion medicine physicians and BSH guidelines | 1 query | ChatGPT3.5:<br>Completeness (2/3)<br>Concordance (3.5/5)<br>ChatGPT4:<br>Completeness (2.25/3)<br>Concordance (3/5)<br>Google Bard:<br>Completeness (1.5/3)<br>Concordance (3/5)<br>Microsoft Bing:<br>Completeness (1.75/3)<br>Concordance (3.75/5) |

**Table 3.**
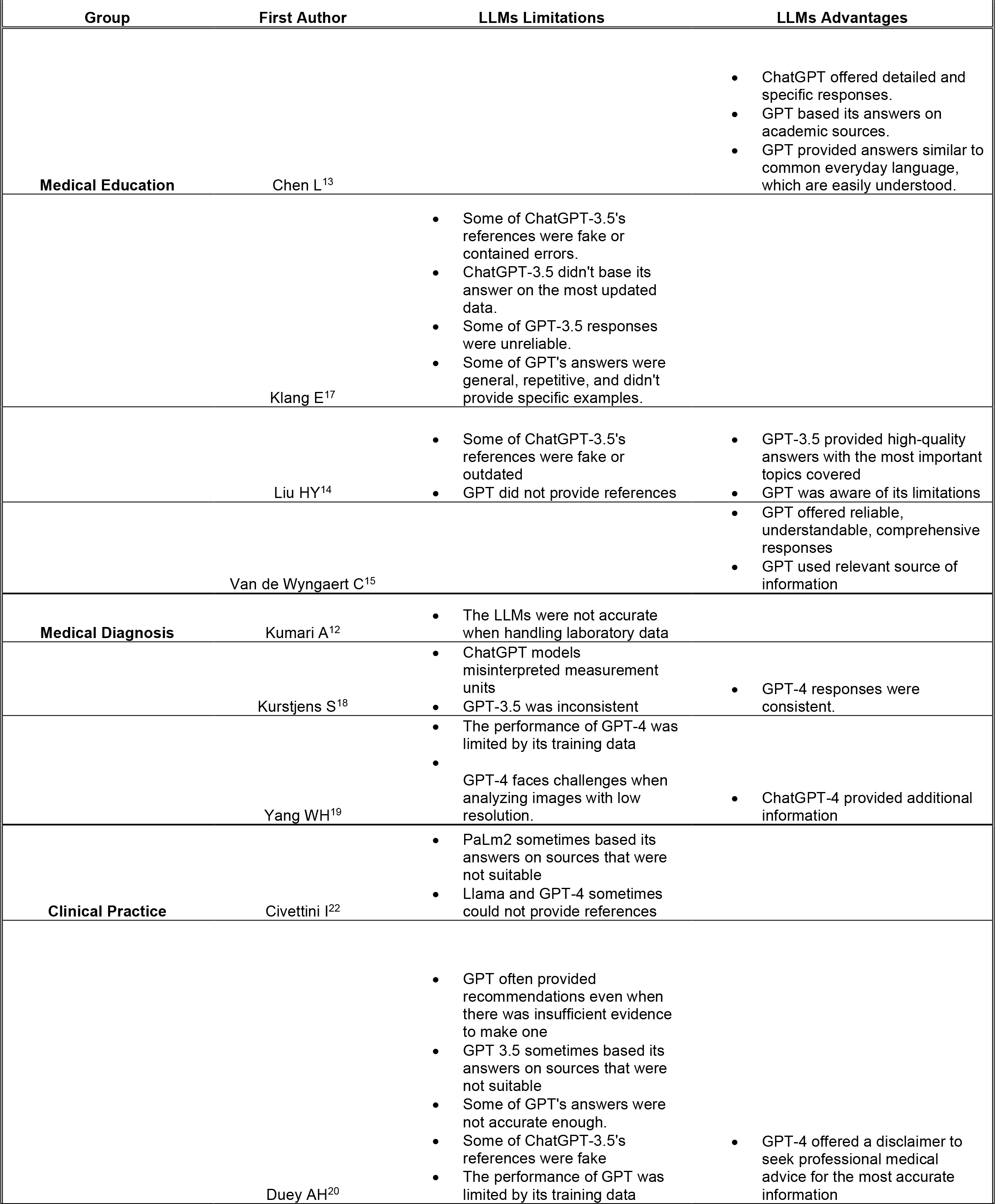
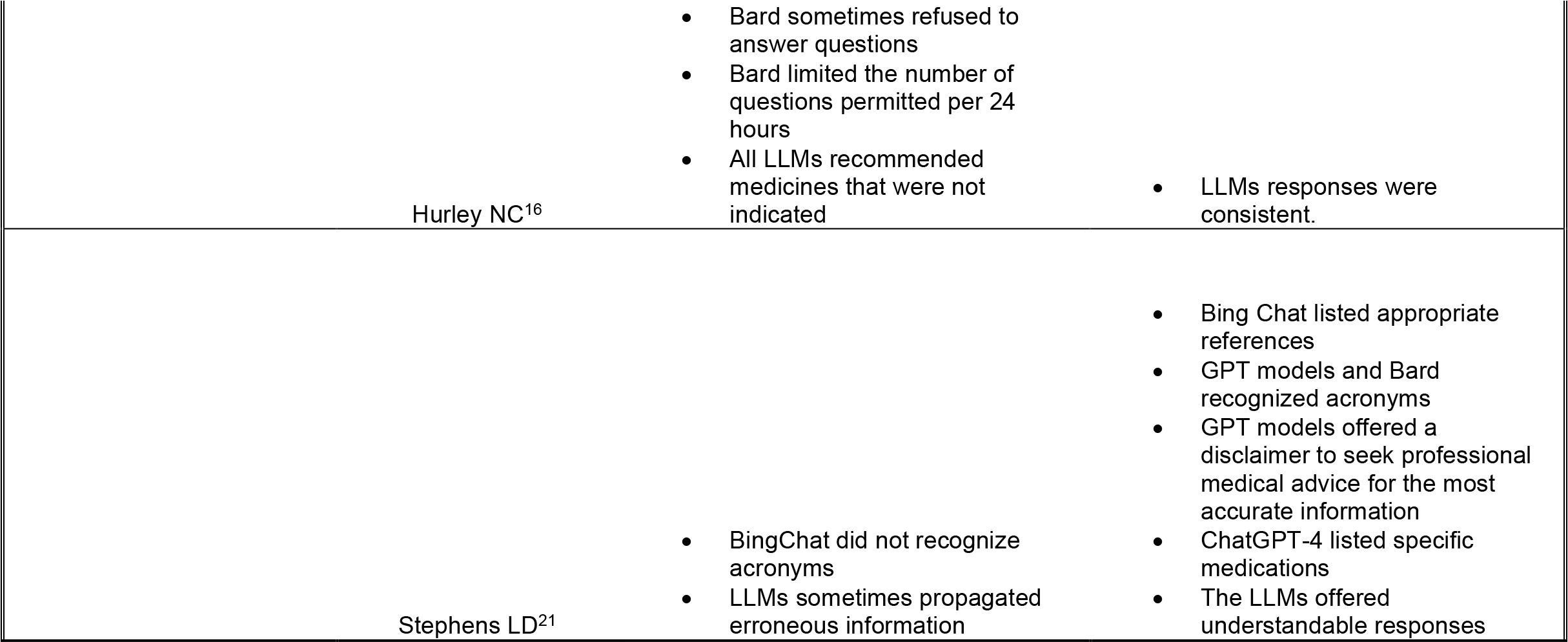
Limitations and Advantages of LLMs in the reviewed articles.

| Group | First Author | LLMs Limitations | LLMs Advantages |
| --- | --- | --- | --- |
| Medical Education | Chen L<sup>13</sup> |  | • ChatGPT offered detailed and specific responses.<br>• GPT based its answers on academic sources.<br>• GPT provided answers similar to common everyday language, which are easily understood. |
|  | Klang E<sup>17</sup> | • Some of ChatGPT-3.5's references were fake or contained errors.<br>• ChatGPT-3.5 didn't base its answer on the most updated data.<br>• Some of GPT-3.5 responses were unreliable.<br>• Some of GPT's answers were general, repetitive, and didn't provide specific examples. |  |
|  | Liu HY<sup>14</sup> | • Some of ChatGPT-3.5's references were fake or outdated<br>• GPT did not provide references | • GPT-3.5 provided high-quality answers with the most important topics covered<br>• GPT was aware of its limitations |
|  | Van de Wyngaert C<sup>15</sup> |  | • GPT offered reliable, understandable, comprehensive responses<br>• GPT used relevant source of information |
| Medical Diagnosis | Kumari A<sup>12</sup> | • The LLMs were not accurate when handling laboratory data |  |
|  | Kurstjens S<sup>18</sup> | • ChatGPT models misinterpreted measurement units<br>• GPT-3.5 was inconsistent | • GPT-4 responses were consistent. |
|  | Yang WH<sup>19</sup> | • The performance of GPT-4 was limited by its training data<br>• GPT-4 faces challenges when analyzing images with low resolution. | • ChatGPT-4 provided additional information |
| Clinical Practice | Civettini I<sup>22</sup> | • PaLM2 sometimes based its answers on sources that were not suitable<br>• Llama and GPT-4 sometimes could not provide references |  |
|  | Duey AH<sup>20</sup> | • GPT often provided recommendations even when there was insufficient evidence to make one<br>• GPT 3.5 sometimes based its answers on sources that were not suitable<br>• Some of GPT's answers were not accurate enough.<br>• Some of ChatGPT-3.5's references were fake<br>• The performance of GPT was limited by its training data | • GPT-4 offered a disclaimer to seek professional medical advice for the most accurate information |

|  |  |  |
| --- | --- | --- |
| Hurley NC<sup>16</sup> | • Bard sometimes refused to answer questions<br>• Bard limited the number of questions permitted per 24 hours<br>• All LLMs recommended medicines that were not indicated | • LLMs responses were consistent. |
| Stephens LD<sup>21</sup> | • BingChat did not recognize acronyms<br>• LLMs sometimes propagated erroneous information | • Bing Chat listed appropriate references<br>• GPT models and Bard recognized acronyms<br>• GPT models offered a disclaimer to seek professional medical advice for the most accurate information<br>• ChatGPT-4 listed specific medications<br>• The LLMs offered understandable responses |

### Medical Education

Two studies compared the effectiveness of ChatGPT (GPT-3.5) with Google search in answering medical questions on stem cell therapy and breast implant-associated anaplastic large cell lymphoma (BIA-ALCL).^13^ ^14^ Both studies highlighted GPT-3.5’s superiority in propagating professional information.

In another study by Van de Wyngaert C. et al., GPT-3.5’s answers to frequently asked questions about hemophilia were assessed and compared to the answers found on specialized websites. GPT’s responses were more relevant, exhaustive, scientifically valid, and understandable, compared to the websites.^15^

When assessing the performance of LLMs on the Internal Medicine postgraduate knowledge of transfusion practice (BEST-TEST), a multiple-choice test regarding transfusion medicine, GPT-4 performed the best with an average of 87% correct answers, followed by Bard (54%) and GPT-3.5 (44%).^16^

Klang et al. demonstrated GPT-3.5’s utility for academic writing in thrombosis and hemostasis. The model’s limitations highlighted the importance of human judgment and medical expertise in the writing of academic papers.^17^

### Medical Diagnosis

Kumari A. et al. compared the capability of different LLMs in solving hematology cases. GPT-3.5 performed the best, achieving a score of 3.2 out of 5, followed by Google’s Bard with a score of 2.2, and Microsoft’s Bing with 2.0.^12^

In a study that investigated the ability of GPT models to diagnose hemoglobinopathy (Beta heterozygote, 3.7 homozygote, SEA, MED, FIL, HbC or HbE heterozygote) by interpreting patients’ laboratory results, GPT-4 achieved a diagnostic accuracy of 76%, with 97% true positives and 56% true negatives. In the same task, GPT-3.5 achieved an accuracy of 53%, with 75% true positives and 31% true negatives.^18^

Yang et al., evaluated the potential of GPT-4 to assist with blood morphology identification. GPT-4 identified normal blood cells with an accuracy of 88%, exceeding the accuracy of identifying abnormal blood cells at a rate of 54%. Regarding identifying abnormal cells, the accuracy of GPT-4 was slightly higher than that of the manual method, which was 49.5%.^19^

### Clinical practice and therapeutic recommendation

Duey et al. compared the recommendations of GPT-3.5 and GPT4 on thromboembolic prophylaxis in spine surgery. They showed that GPT-3.5 had an accuracy rate of 33%, while ChatGPT-4.0 achieved an accuracy of 92%.^20^

The performance of GPT models and Google Bard in answering common questions related to clinical transfusion practice was better with explicitly phrased questions compared to those with implicit, more "realistic" phrasing. GPT-4 performed best with a score of 83% on implicit questions and 100% on explicit questions. GPT-3.5 performance was the most affected by question phrasing, with a score of 17% on the implicit questions and 83% on explicit questions.^16^

Stephens et al. evaluated the quality of indications for irradiated blood components for TA-GVHD prevention, as provided by GPT models, Google Bard, and Microsoft Bing. Bing’s responses were rated the most accurate, with a grade of 3.75 out of 5, followed by GPT-3.5 with 3.5, GPT-4 with 3, and Bard with 3. GPT-4’s responses were considered the most complete, receiving a completeness grade of 2.25 out of 3, followed by GPT-3.5 with 2, Bing with 1.75, and Bard with 1.5.^21^

A study by Hurley NC. et al. evaluated the recommendations of GPT models and Google Bard, regarding the necessity for red blood cell transfusion, based on short case presentations. GPT-4 demonstrated the highest performance, with a correct result rate of 91%. Bard performed least well, with a correct result rate of 45.5%.^16^ When assessing the capabilities of GPT-4, PaLm2, Llama2-13b, and Llama2-70b in making complex decisions in hematopoietic stem cell transplantation, only 58.8% of the LLMs’ answers matched the experts’ positions.^22^

#### Benefits and limitations of LLMs in hematology

From the included studies, several strengths and limitations of LLMs in hematology were identified **(Table 3**, **Figure 5)**.

**Figure 1.**
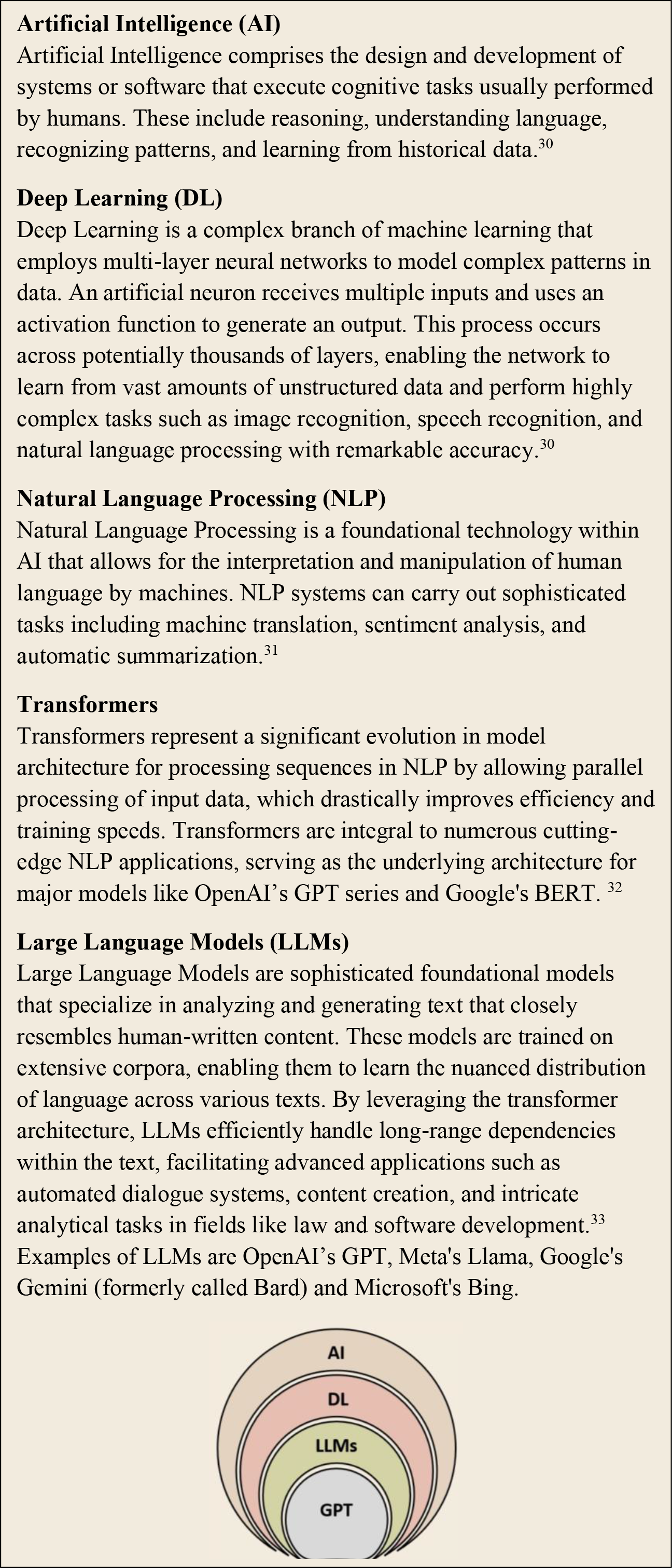
An Overview of Foundational Terms in the field of LLMs and a hierarchy diagram.

**Figure 2.**
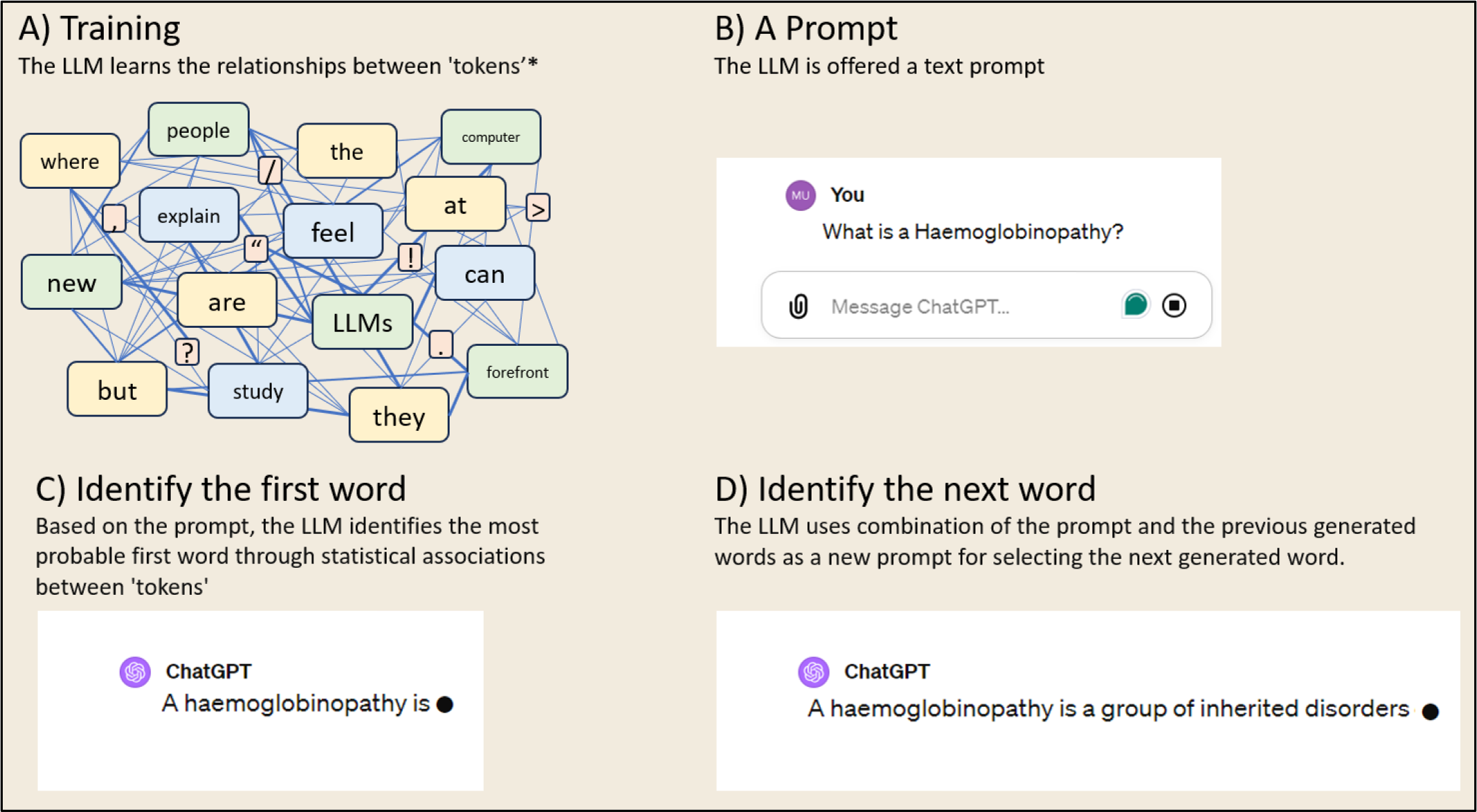
Diagram of the way LLMs are developed

**Figure 3.**
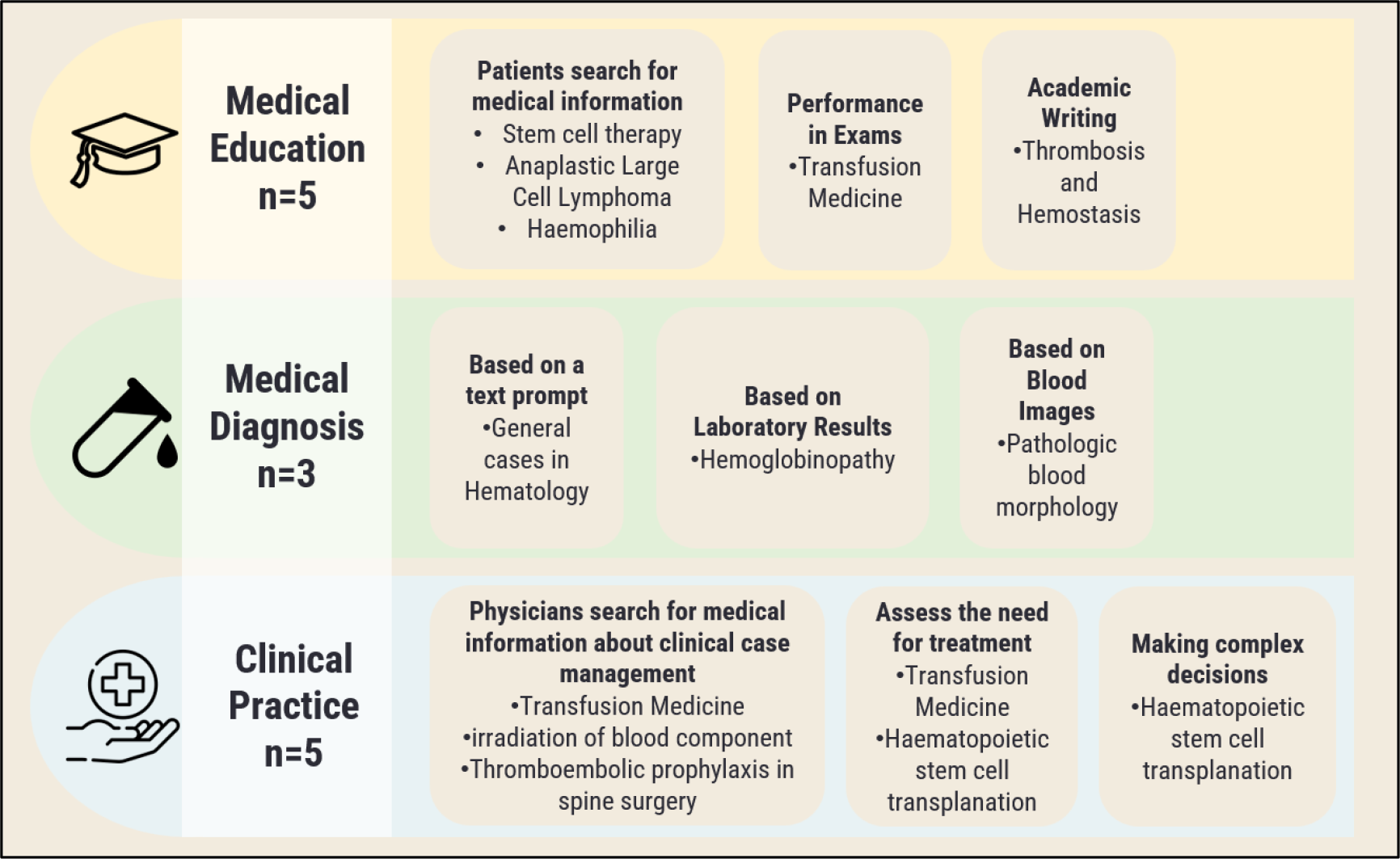
Applications of LLMs in Hematology in the Articles Reviewed

**Figure 4.**
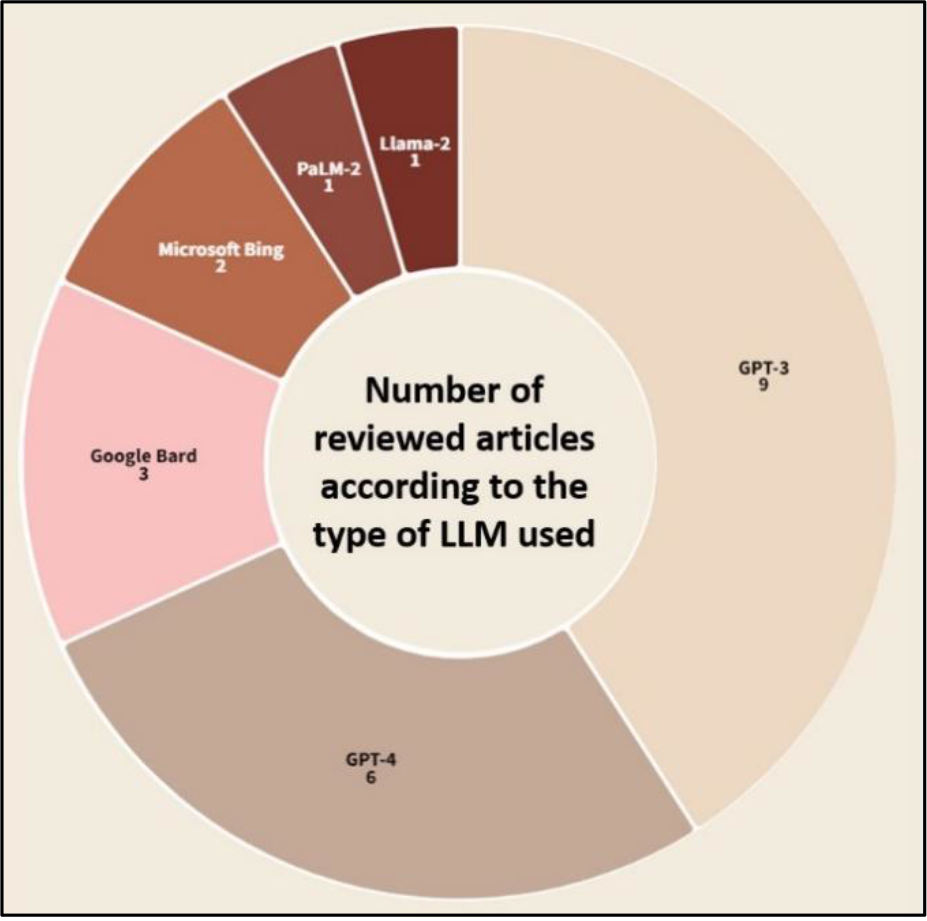
Number of reviewed articles according to the type of LLM used.

**Figure 5.**
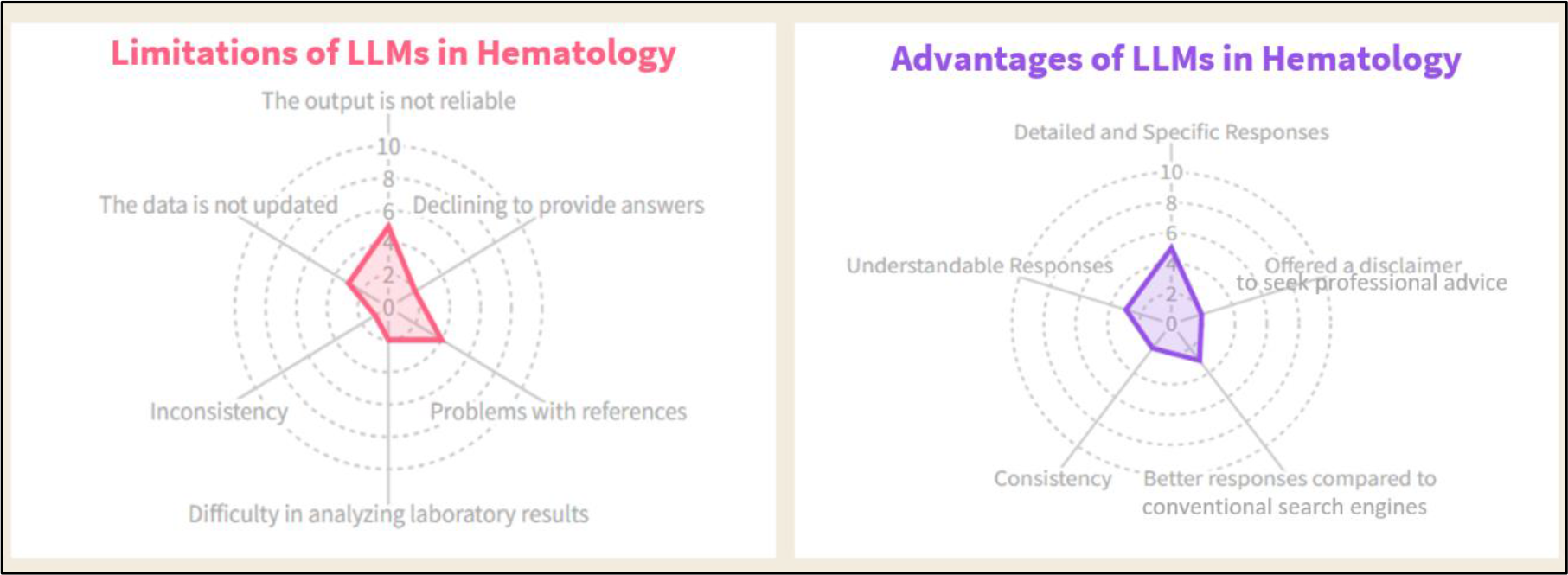
Main Limitations and Advantages of LLMs in Hematology in the Articles Reviewed

Opinions regarding the consistency of GPT models in the field of hematology were divided. The study by Kurstjens S. et al.^18^ described ChatGPT as an inconsistent model (choosing the same answer 80% of the time), while referring to GPT-4 as a very consistent one (choosing the same answer 98.3% of the time). In contrast, Hurley et al.^16^ characterize three LLMs as consistent models, with Bard, GPT-3.5, and GPT-4 choosing the same answer 87%, 99%, and 97% of the time, respectively. The discrepancy highlights the debate within the field regarding the reliability of these models’ outputs.

Some studies highlighted issues with the references used by the LLMs. Three studies reported that some of GPT-3.5’s references were fake, outdated, or contained errors.^14^ ^17^ ^20^ Another study, by Civettini et al., indicated that in some cases, Llama-2 and GPT-4 could not provide references.^22^ Duey et al. observed that GPT-4 did not explicitly cite any sources in its responses, thus avoiding the display of problematic references.^20^ Nonetheless, it is essential to observe that Bing Chat provided appropriate references in the study by Stephens et al. .^21^

Three studies noted limitations in the performance of GPT models due to the large datasets used to train them, which are not updated enough. One focused on GPT-3.5,^17^ another on GPT-4,^19^ a third on both.^20^ Van de Wyngaert et al. also mentioned the potential impact of this issue on ChatGPT’s performance.^15^

Concerns regarding the reliability of the data were raised. GPT-3.5^20^ and PaLm2^22^ might derive their answers from sources that are not suitable, leading to potentially incorrect responses. For example, PaLm2 consistently based its responses on UpToDate (uptodate.com) information on stem cell transplantation in acute myeloid leukemia, not only when asked about acute myeloid leukemia but also when asked about acute lymphoblastic leukemia.

Other studies pointed out that some of LLMs’ answers are not accurate enough.^17^ ^20^ ^21^ For instance, LLMs recommended medications that were not indicated,^16^ and GPT models offered recommendations even when there was insufficient evidence.^20^ However, additional research suggests that GPT’s responses are reliable and based on academic sources.^13^ ^15^

LLMs also encountered some problems when dealing with laboratory data.^12^ ^18^ For example, ChatGPT interpreted hemoglobin results, expressed in nmol/L, as if the concentration was in g/dL.^18^

An important strength that was observed in the reviewed articles is that LLMs offered detailed and specific responses,^13^ ^14^ ^15^ ^19^ ^21^ written in a manner similar to common everyday language.^13^ Additionally, GPT models sometimes included a disclaimer advising users to seek professional medical advice for the most accurate information.^20^ ^21^

## DISCUSSION

This systematic review examines the use of Large Language Models (LLMs) such as OpenAI’s ChatGPT and Google’s Bard within the field of Hematology. It highlights the potential and the limitations of these models in enhancing medical education, advancing diagnostic accuracy, and shaping clinical practice within the field.

### Evaluation of LLMs in Medical Education, Diagnosis and Clinical Decision-making

The studies reviewed demonstrate that LLMs can surpass traditional electronic methods in delivering medical education to both patients and physicians. For example, ChatGPT provided responses that were more relevant and easier to understand than those of standard search engines and specialized medical websites.^13^ ^14^ ^15^ Patient interaction with chatbot can help patients understand their illness and treatment options. It can also serve as a translator for medical terminology used by physicians. This capability suggests that LLMs could play a pivotal role in democratizing access to reliable medical information, thereby potentially transforming medical education by making high-quality knowledge more accessible.

In diagnostic applications, the performance of LLMs varied, with some models achieving diagnostic accuracies that rival traditional methods.^19^ This variance underscores the importance of ongoing model training and validation to ensure that these tools provide accurate and reliable diagnostics.

The application of LLMs in clinical settings has shown that while they can offer accurate recommendations for treatment and diagnosis, their reliability can be inconsistent.^16^ ^20^ ^21^ ^22^ This inconsistency is particularly concerning given the high stakes of medical decision-making. For instance, the studies highlighted issues with the models’ references^14^ ^17^ ^20^ ^22^ and the reliability of their training data,^20^ ^22^ which could lead to incorrect or outdated medical advice.

Moreover, the variation in the performance of different LLMs in handling implicit versus explicit queries raises concerns about their practicality in real-world clinical environments, where queries may not always be clearly formulated.^16^ This limitation could affect the models’ utility in emergency settings or complex cases where nuanced understanding and rapid decision-making are crucial.

### The Potential of LLMs in Hematology

While LLMs are often praised for their potential to streamline operations and enhance decision-making, it is crucial to acknowledge the implications of their integration into healthcare.^23^ ^24^ The reliance on AI-generated knowledge could shift the focus of medical authority from trained professionals to AI systems, altering the power dynamics in healthcare settings. Such a shift could exacerbate existing challenges related to trust and accountability in medical practice.

Additionally, the adoption of LLMs might lead to a homogenization of medical knowledge, as these models are trained primarily on existing datasets that may not fully capture the diversity of patient experiences or the complexities of rare conditions.^25^ This could stifle innovation in medical thinking and reduce the personalized nature of patient care, potentially leading to a one-size-fits-all approach that neglects individual patient needs and contexts.

Another important point is the critical understanding of how chatbots function, as the design of user prompts greatly influences the responses they generate. In research settings, prompts are meticulously crafted to ensure precision and relevance; however, this level of detail often diminishes in real-world applications. To secure accurate, non-generic responses, users must clearly specify their audience and the context of their questions. Considering the challenges associated with creating effective prompts, customizing LLM to meet the diverse needs of hematology patients is extremely important.

### Future Directions

The applications of LLMs in hematology discussed in this review are currently in their early stages, and the full spectrum of their potential remains largely untapped. There is much to discover about what LLMs can offer to the field. There are many more subareas of hematology, such as leukemias, coagulation disorders, and myelodysplastic syndromes, that have not yet been studied using LLMs. Additionally, there are many more tasks where LLMs could make significant contributions including, identifying adverse events, enriching risk prediction models, enhancing patient management, as well as improving administrative processes and compliance to guidelines.^26^

To harness the benefits of LLMs while mitigating the risks, future research should focus on enhancing the transparency and accountability of these models. Developing standards for the ethical use of AI in medicine, improving the diversity and reliability of training datasets, and implementing robust validation processes are critical steps toward responsible integration.

Furthermore, fostering a collaborative environment where AI complements rather than replaces human expertise could help maintain the essential role of medical professionals’ judgment. Integrating LLMs into clinical practice should not diminish the value of human expertise but rather augment it, ensuring that medical care remains compassionate, individualized, and informed by both human empathy and AI’s analytical capabilities.

This systematic review encounters several limitations. The small number of studies included restricts the ability to draw broad conclusions. Variability in study tasks and methods hinders the execution of a meta-analysis. Additionally, as the investigation of LLMs in this field is relatively recent, the long-term impacts remain uncertain. Concerns regarding the training data—specifically its lack of diversity and inherent biases—could compromise the reliability of AI-generated recommendations in clinical practice.

Furthermore, the continuous improvement in LLM technology means that the performance of an LLM noted in a reported study may not accurately reflect its current capabilities. For example, in December 2023, Google made some significant improvements to Bard, including renaming it to Gemini.^27^ Moreover,

In November 2023, Elon Musk’s xAI introduced a new AI model named Grok. The development team highlights that Grok distinguishes itself from existing LLMs by its ability to generate responses based on ’real-time knowledge,’ a feature which provides users with the most updated information.^28^

In conclusion, While LLMs offer substantial opportunities for advancing hematology, it is important to recognize that the field is still in its early stages, not yet addressing many diseases and potential tasks. Moreover, its development is limited by variability, reliability issues, and inaccuracies. As such, integrating LLMs into clinical practice must be navigated with careful consideration of both their potential and their pitfalls.

Balancing innovation with caution will be key to realizing the benefits of LLMs without compromising the integrity and human-centered nature of medical care.

## Supporting information

Supplementary Materials ("Detailed Search Terms")

PRISMA

## Data Availability

All data produced in the present work are contained in the manuscript

## REFERENCES

1. Meyrowitsch DW, Jensen AK, Sørensen JB, Varga T V. AI chatbots and (mis)information in public health: impact on vulnerable communities. Front Public Health. 2023;11:1226776. doi:10.3389/fpubh.2023.1226776

2. Clusmann J, Kolbinger FR, Muti HS, et al. The future landscape of large language models in medicine. Communications Medicine. 2023;3(1):141. doi:10.1038/s43856-023-00370-1

3. Abd-Alrazaq A, AlSaad R, Alhuwail D, et al. Large Language Models in Medical Education: Opportunities, Challenges, and Future Directions. JMIR Med Educ. 2023;9:e48291. doi:10.2196/48291

4. Yang X, Chen A, PourNejatian N, et al. A large language model for electronic health records. NPJ Digit Med. 2022;5(1):194. doi:10.1038/s41746-022-00742-2

5. Cascella M, Semeraro F, Montomoli J, Bellini V, Piazza O, Bignami E. The Breakthrough of Large Language Models Release for Medical Applications: 1-Year Timeline and Perspectives. J Med Syst. 2024;48(1):22. doi:10.1007/s10916-024-02045-3

6. Deng J, Zubair A, Park YJ. Limitations of large language models in medical applications. Postgrad Med J. 2023;99(1178):1298-1299. doi:10.1093/postmj/qgad069

7. Ullah E, Parwani A, Baig MM, Singh R. Challenges and barriers of using large language models (LLM) such as ChatGPT for diagnostic medicine with a focus on digital pathology - a recent scoping review. Diagn Pathol. 2024;19(1):43. doi:10.1186/s13000-024-01464-7

8. Moher D. Preferred Reporting Items for Systematic Reviews and Meta-Analyses: The PRISMA Statement. Ann Intern Med. 2009;151(4):264. doi:10.7326/0003-4819-151-4-200908180-00135

9. McInnes MDF, Moher D, Thombs BD, et al. Preferred Reporting Items for a Systematic Review and Meta-analysis of Diagnostic Test Accuracy Studies. JAMA. 2018;319(4):388. doi:10.1001/jama.2017.19163

10. Moons KGM, de Groot JAH, Bouwmeester W, et al. Critical Appraisal and Data Extraction for Systematic Reviews of Prediction Modelling Studies: The CHARMS Checklist. PLoS Med. 2014;11(10):e1001744. doi:10.1371/journal.pmed.1001744

11. Whiting PF. QUADAS-2: A Revised Tool for the Quality Assessment of Diagnostic Accuracy Studies. Ann Intern Med. 2011;155(8):529. doi:10.7326/0003-4819-155-8-201110180-00009

12. Kumari A, Kumari A, Singh A, et al. Large Language Models in Hematology Case Solving: A Comparative Study of ChatGPT-3.5, Google Bard, and Microsoft Bing. Cureus. 2023;15(8):e43861. doi:10.7759/cureus.43861

13. Chen L, Li H, Su Y, et al. Using A Google Web Search Analysis to Assess the Utility of ChatGPT in Stem Cell Therapy. Stem Cells Transl Med. 2024;13(1):60–68. doi:10.1093/stcltm/szad074

14. Liu HY, Alessandri Bonetti M, De Lorenzi F, Gimbel ML, Nguyen VT, Egro FM. Consulting the Digital Doctor: Google Versus ChatGPT as Sources of Information on Breast Implant-Associated Anaplastic Large Cell Lymphoma and Breast Implant Illness. Aesthetic Plast Surg. 2024;48(4):590–607. doi:10.1007/s00266-023- 03713-4

15. Van de Wyngaert C, Iarossi M, Hermans C. How good does ChatGPT answer frequently asked questions about haemophilia? Haemophilia. 2023;29(6):1646–1648. doi:10.1111/hae.14858

16. Hurley NC, Schroeder KM, Hess AS. Would doctors dream of electric blood bankers? Large language model-based artificial intelligence performs well in many aspects of transfusion medicine. Transfusion (Paris). 2023;63(10):1833–1840. doi:10.1111/trf.17526

17. Klang E, Levy-Mendelovich S. Evaluation of OpenAI’s large language model as a new tool for writing papers in the field of thrombosis and hemostasis. J Thromb Haemost. 2023;21(4):1055–1058. doi:10.1016/j.jtha.2023.01.011

18. Kurstjens S, Schipper A, Krabbe J, Kusters R. Predicting hemoglobinopathies using ChatGPT. Clin Chem Lab Med. 2024;62(3):e59–e61. doi:10.1515/cclm-2023-0885

19. Yang WH, Yang YJ, Chen TJ. ChatGPT’s innovative application in blood morphology recognition. J Chin Med Assoc. 2024;87(4):428–433. doi:10.1097/JCMA.0000000000001071

20. Duey AH, Nietsch KS, Zaidat B, et al. Thromboembolic prophylaxis in spine surgery: an analysis of ChatGPT recommendations. Spine J. 2023;23(11):1684–1691. doi:10.1016/j.spinee.2023.07.015

21. Stephens LD, Jacobs JW, Adkins BD, Booth GS. Battle of the (Chat)Bots: Comparing Large Language Models to Practice Guidelines for Transfusion-Associated Graft-Versus-Host Disease Prevention. Transfus Med Rev. 2023;37(3):150753. doi:10.1016/j.tmrv.2023.150753

22. Civettini I, Zappaterra A, Granelli BM, et al. Evaluating the performance of large language models in haematopoietic stem cell transplantation decision-making. Br J Haematol. 2024;204(4):1523–1528. doi:10.1111/bjh.19200

23. Omiye JA, Gui H, Rezaei SJ, Zou J, Daneshjou R. Large Language Models in Medicine: The Potentials and Pitfalls. Ann Intern Med. 2024;177(2):210–220. doi:10.7326/M23-2772

24. Thirunavukarasu AJ, Ting DSJ, Elangovan K, Gutierrez L, Tan TF, Ting DSW. Large language models in medicine. Nat Med. 2023;29(8):1930–1940. doi:10.1038/s41591-023-02448-8

25. Singhal K, Azizi S, Tu T, et al. Large language models encode clinical knowledge. Nature. 2023;620(7972):172-180. doi:10.1038/s41586-023-06291-2

26. Boonstra MJ, Weissenbacher D, Moore JH, Gonzalez-Hernandez G, Asselbergs FW. Artificial intelligence: revolutionizing cardiology with large language models. Eur Heart J. 2024;45(5):332–345. doi:10.1093/eurheartj/ehad838

27. 27. Team G (2024) Bard becomes Gemini: try Ultra 1.0 and a new mobile app today. Google, Inc. https://blog.google/products/gemini/bard-gemini-advanced-app/.

28. Announcing grok. Announcing grok. Accessed December 21, 2023. https://x.ai/.

29. Naveed H, Khan AU, Qiu S, et al. A Comprehensive Overview of Large Language Models. Published online July 12, 2023.

30. Soffer S, Ben-Cohen A, Shimon O, Amitai MM, Greenspan H, Klang E. Convolutional Neural Networks for Radiologic Images: A Radiologist’s Guide. Radiology. 2019;290(3):590–606. doi:10.1148/radiol.2018180547

31. Sorin V, Barash Y, Konen E, Klang E. Deep Learning for Natural Language Processing in Radiology- Fundamentals and a Systematic Review. J Am Coll Radiol. 2020;17(5):639–648. doi:10.1016/j.jacr.2019.12.026

32. Soffer S, Glicksberg BS, Zimlichman E, Klang E. BERT for the Processing of Radiological Reports: An Attention-based Natural Language Processing Algorithm. Acad Radiol. 2022;29(4):634–635. doi:10.1016/j.acra.2021.03.036

33. Almarie B, Teixeira PEP, Pacheco-Barrios K, Rossetti CA, Fregni F. Editorial - The Use of Large Language Models in Science: Opportunities and Challenges. Princ Pract Clin Res. 2023;9(1):1–4. doi:10.21801/ppcrj.2023.91.1

