## Supplementary Materials ("Detailed Search Terms") for "Exploring the role of Large Language Models (LLMs) in hematology: a systematic review of applications, benefits, and limitations"

**Detailed Search Strategies**

**PubMed (PubMed.gov):**

**#1**
(Hematology) OR (Blood Disorders) OR (Blood Diseases) OR (Blood Conditions) OR (Anemia) OR (Sickle Cell Disease) OR (Thalassemia) OR (Leukemia) OR (Lymphoma) OR (Myeloma) OR (Hodgkin's Lymphoma) OR (Non-Hodgkin's Lymphoma) OR (Blood Cancer) OR (Hematologic Neoplasms) OR (Platelet Disorders) OR (Hemophilia) OR (Von Willebrand Disease) OR (Coagulation Disorders) OR (Bleeding Disorders) OR (Thrombosis) OR (Deep vein thrombosis) OR (Pulmonary embolism) OR (Iron Deficiency) OR (Hemochromatosis) OR (Polycythemia Vera) OR (Myelofibrosis) OR (Essential thrombocytosis) OR (Blood Transfusion) OR (Stem Cell Transplantation) OR (Bone Marrow Transplantation) OR (Diagnostic Hematology) OR (Laboratory Hematology) OR (Hematopoiesis) OR (Erythropoiesis) OR (Hematologic Tests) OR (Complete Blood Count) OR (Hemic and Lymphatic Diseases) OR (hemostasis)

**#2**
("ChatGPT") OR ("large language models") OR ("OpenAI") OR ("Microsoft bing") OR ("google bard") OR ("google gemini")

**#3**
(#1 AND #2) AND English **Filters:** from 2023 – 2024

**Scopus (Scopus.com):**

**#1
( "Hematology" ) OR ( "Blood Disorders" ) OR ( "Blood Diseases" ) OR ( "Blood Conditions" ) OR ( "Anemia" ) OR ( "Sickle Cell Disease" ) OR ( "Thalassemia" ) OR ( "Leukemia" ) OR ( "Lymphoma" ) OR ( "Myeloma" ) OR ( "Hodgkin&apos;s Lymphoma" ) OR ( "Non-Hodgkin Lymphoma" ) OR ( "Blood Cancer" ) OR ( "Hematologic Neoplasms" ) OR ( "Platelet Disorders" ) OR ( "Hemophilia" ) OR ( "Von Willebrand Disease" ) OR ( "Coagulation Disorders" ) OR ( "Bleeding Disorders" ) OR ( thrombosis ) OR ( "Deep vein thrombosis" ) OR ( "Pulmonary embolism" ) OR ( "Iron Deficiency" ) OR ( hemochromatosis ) OR ( "Polycythemia Vera" ) OR ( myelofibrosis ) OR ( "Essential thrombocytosis" ) OR ( "Blood Transfusion" ) OR ( "Stem Cell Transplantation" ) OR ( "Bone Marrow Transplantation" ) OR ( "Diagnostic Hematology" ) OR ( "Laboratory Hematology" ) OR ( hematopoiesis ) OR ( erythropoiesis ) OR ( "Hematologic Tests" ) OR ( "Complete Blood Count" ) OR ( "Hemic and Lymphatic Diseases" ) OR ("hemostasis")**

**#2
( "ChatGPT" ) OR ( "large language models" ) OR ( "OpenAI" ) OR ( "Microsoft bing" ) OR ( "google bard" ) OR ( "google gemini")**

**#3**
(#1 AND #2) AND **PUBYEAR > 2022 AND ( LIMIT-TO ( DOCTYPE , "ar" ) ) AND ( LIMIT-TO ( SUBJAREA , "MEDI" ) ) AND ( LIMIT-TO ( LANGUAGE , "English" ) )**

**Web of Science (Thomson Reuters):**

**#1
ALL=(Hematology)) OR ALL=(Blood Disorders)) OR ALL=(Blood Diseases)) OR ALL=(Blood Conditions)) OR ALL=(Anemia)) OR ALL=(Sickle Cell Disease)) OR ALL=(Thalassemia)) OR ALL=(Leukemia)) OR ALL=(Lymphoma)) OR ALL=(Myeloma)) OR ALL=(Hodgkin's Lymphoma)) OR ALL=(Non-Hodgkin's Lymphoma)) OR ALL=(Blood Cancer)) OR ALL=(Hematologic Neoplasms)) OR ALL=(Platelet Disorders)) OR ALL=(Hemophilia)) OR ALL=(Von Willebrand Disease)) OR ALL=(Coagulation Disorders)) OR ALL=(Bleeding Disorders)) OR ALL=(Thrombosis)) OR ALL=(Deep vein thrombosis)) OR ALL=(Pulmonary embolism)) OR ALL=(Iron Deficiency)) OR ALL=(Hemochromatosis)) OR ALL=(Polycythemia Vera)) OR ALL=(Myelofibrosis)) OR ALL=(Essential thrombocytosis)) OR ALL=(Blood Transfusion)) OR ALL=(Stem Cell Transplantation)) OR ALL=(Bone Marrow Transplantation)) OR ALL=(Diagnostic Hematology)) OR ALL=(Laboratory Hematology)) OR ALL=(Hematopoiesis)) OR ALL=(Erythropoiesis)) OR ALL=(Hematologic Tests)) OR ALL=(Complete Blood Count)) OR ALL=(Hemic and Lymphatic Diseases)) OR ALL=(hemostasis)**

**#2
ALL=(ChatGPT)) OR ALL=(large language models)) OR ALL=(OpenAI)) OR ALL=(Microsoft bing)) OR ALL=(google bard)) OR ALL=(google gemini)**

**#3**
(#1 AND #2)
