## Supplementary Figure 1 for "Exploring the role of Large Language Models (LLMs) in hematology: a systematic review of applications, benefits, and limitations"

**PRISMA flow diagram**

**PRISMA flow diagram**

Records identified through Pubmed (n = 95)

Records identified through
Web of Science (n = 120)

Records identified through
Scopus (n = 110)

**Identification**

Records identified through all databases searching

(n = 325)

Duplicate records removed *before screening*

(n = 89)

Records screened

(n = 236)

Records excluded:
Unrelated to subject (n = 221)

Reports excluded:

Not an original article (n = 2)

Was written by the assistance of LLMs (n = 2)

**Screening**

Reports assessed for eligibility

(n = 15)

**Included**

Studies included in review

(n = 11)

**Supplementary Figure 1.** Flow diagram of the search and inclusion process.
