## Supplementary Table 1 for "Exploring the role of Large Language Models (LLMs) in hematology: a systematic review of applications, benefits, and limitations"

**QUADAS-2 risk of bias**

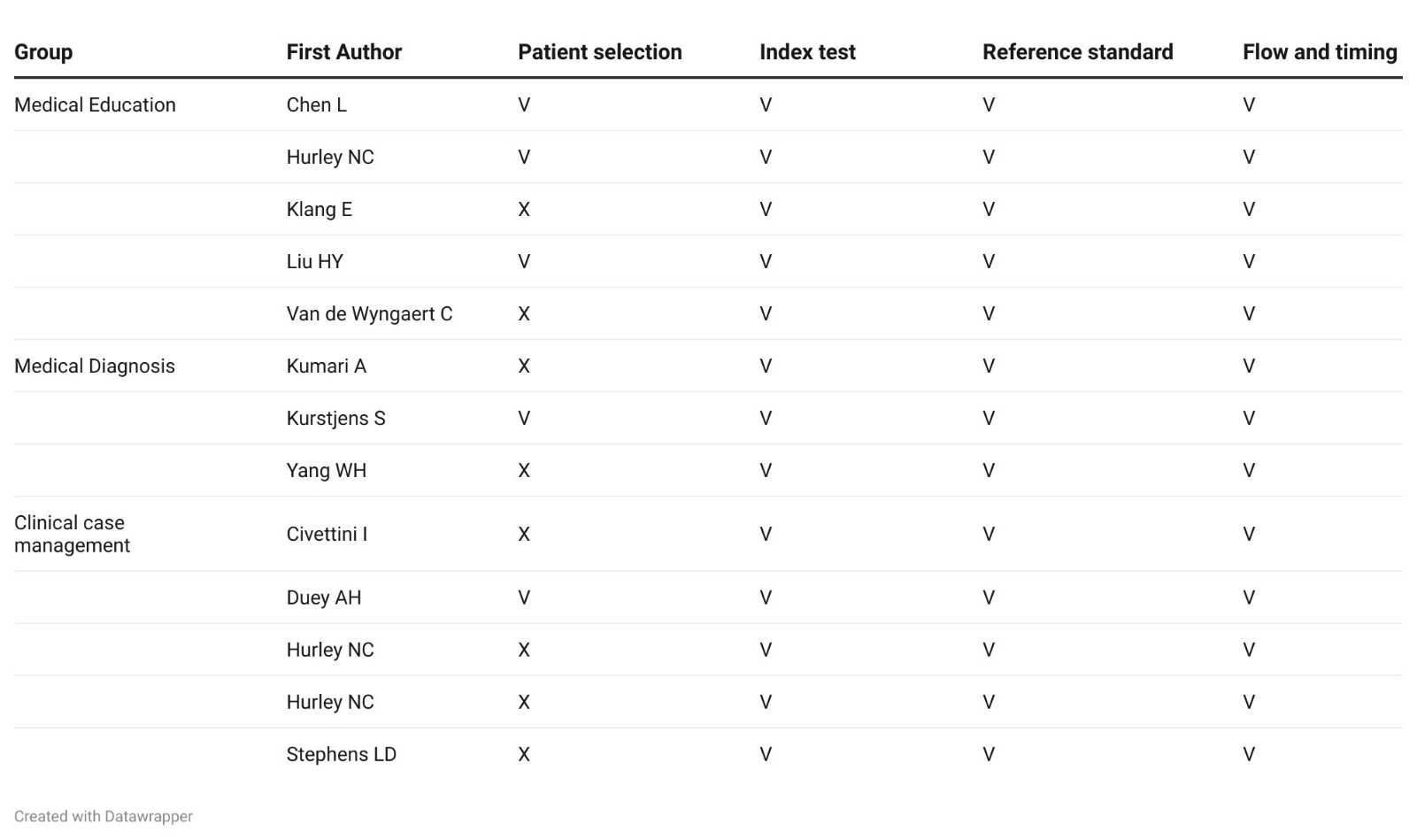

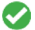

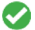

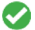

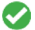

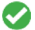

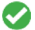

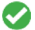

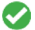

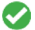

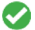

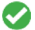

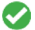

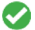

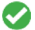

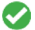

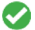

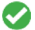

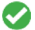

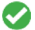

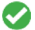

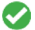

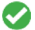

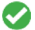

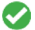

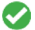

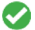

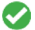

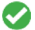

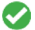

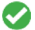

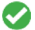

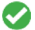

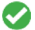

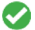

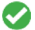

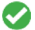

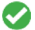

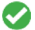

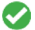

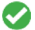

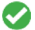

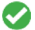

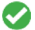

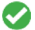

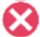

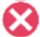

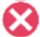

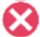

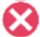

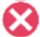

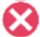

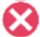

**Supplementary Table 1.** QUADAS-2 risk of bias assessment per clinical application
